# Evaluating MRI-Confirmed Relapses as a Novel Primary Endpoint in Multiple Sclerosis Trials

**DOI:** 10.1101/2025.10.10.25337738

**Authors:** Antoine Gavoille, Stanislas Demuth, Mikaïl Nourredine, Igor Faddeenkov, Fabien Rollot, Romain Casey, Anne Kerbrat, Emmanuelle Le Page, Kevin Bigaut, Guillaume Mathey, Laure Michel, Jonathan Ciron, Aurélie Ruet, Elisabeth Maillart, Pierre Labauge, Hélène Zephir, Caroline Papeix, Gilles Defer, Christine Lebrun-Frenay, Thibault Moreau, Eric Berger, Bruno Stankoff, Pierre Clavelou, Eric Thouvenot, Olivier Heinzlef, Jean Pelletier, Abdullatif Al-Khedr, Olivier Casez, Bertrand Bourre, Abir Wahab, Laurent Magy, Jean-Philippe Camdessanché, Inès Doghri, Solène Moulin, Mariana Sarov-Rivière, Karolina Hankiewicz, Amélie Dos Santos, Corinne Pottier, Chantal Nifle, Eric Manchon, Maia Tchikviladze, Béatrice Baciotti, Marianne Payet, Laura Luciani, Sandrine Wiertlewski, Jérôme De Sèze, Pierre-Antoine Gourraud, Gilles Edan, Sandra Vukusic, David-Axel Laplaud, OFSEP and PRIMUS investigators

## Abstract

**Background:** Clinically defined relapses are the traditional primary endpoint of randomized control trials (RCTs) in multiple sclerosis (MS), yet a substantial proportion lack new inflammatory lesions. Confirming relapses with brain and spinal cord MRI to distinguish relapses with active MRI (RAM) from acute clinical events with stable MRI (ACES) may provide a more sensitive primary outcome for future trials.

**Objective:** To estimate RAM and ACES rates in MS trials and evaluate the impact on statistical power of using RAM.

**Methods:** We used two approaches: an aggregated data (AD) approach, combining population-level data from RCTs with observational data from the French MS registry, and an individual patient data (IPD) approach from the PRIMUS platform. Trials were selected if they evaluated DMTs sufficiently represented in the OFSEP ancillary study or were available in PRIMUS. Eleven pivotal RCTs were included, evaluating natalizumab, cladribine, dimethyl fumarate, teriflunomide, fingolimod, or ocrelizumab; 7 were analyzed with AD only, 1 with IPD only, and 3 with both. For the AD approach, population-level characteristics were extracted from published reports; expected RAM probabilities were then derived from a RAM model fitted on OFSEP observational data, applied to each RCT arm population. For the IPD approach, clinically defined relapses were directly classified as RAM/ACES according to radiological activity on subsequent brain MRI. Main outcomes were the treatment effect on RAM and ACES rates, compared with the effect on clinically defined relapses.

**Results:** Across 11 RCTs, treatment effects were consistently equal or greater for RAM than for clinically defined relapses, with both AD and IPD approaches. No DMT significantly reduced ACES rates, which remained stable at approximately 0.08 events/year across arms. The IPD approach yielded systematically lower RAM probabilities than the AD approach.

Using RAM as the endpoint improved statistical power in most scenarios: *e.g.* a trial with annualized relapse rates of 0.15/year (active arm) vs 0.30 (control arm) requires one-third fewer participants.

**Conclusion:** Adopting RAM as the primary outcome could substantially enhance the power of future MS trials and better target the effect of treatment on inflammatory activity.

## INTRODUCTION

In multiple sclerosis (MS), relapses are defined exclusively by clinical criteria: a monophasic episode with patient-reported symptoms and objective neurological findings typical of MS, developing acutely or subacutely, lasting more than 24 hours, without fever or infection.^1^ For decades, such clinically defined relapses have been the primary endpoint in randomized clinical trials (RCTs) to assess the efficacy of disease-modifying therapies (DMTs) in patients with relapsing MS.^2–11^ However, the decline in annualized relapse rate (ARR) in both treated and control arms driven by increasingly effective therapies poses a major methodological challenge for modern trials, as it reduces the power to demonstrate the efficacy of new treatments on inflammatory activity.^12,13^

In a recent proof-of-concept study on observational data from the French MS registry, the *Observatoire Français de la Sclérose en Plaques* (OFSEP), approximately 25% of clinical events qualified as relapses by neurologists were not associated with radiological activity (new/enlarging T2-lesions and/or new gadolinium-enhanced T1-lesions) on a subsequent brain and spinal cord MRI.^14^ We termed these events “acute clinical events with stable MRI” (ACES), as opposed to “relapses with active MRI” (RAM). Furthermore, while DMTs significantly reduces RAM rates, ACES rates remained remarkably stable across all DMT groups, suggesting that ACES may not represent a therapeutic target of current DMTs.

In RCTs, this RAM/ACES distinction has important methodological implications. If a treatment’s goal is to reduce the RAM rate, then it could be beneficial to confirm each suspected clinical relapse with a comprehensive brain and spinal cord MRI. Using a signal-to-noise analogy, the treatment effect on RAM can be seen as the signal of interest, whereas ACES constitute a background noise that dilutes this signal, biasing the treatment effect toward the null.^15^ Therefore, using MRI-confirmed relapses as the primary outcome could improve statistical power in future MS trials, reducing the required sample size or trial duration substantially. Moreover, in inconclusive trials, it could clarify whether results reflect an absence of efficacy or insufficient power caused by a low number of events, as seen recently in the evobrutinib and tolebrutinib trials in relapsing MS.^12,13^

In this study, we estimated RAM and ACES rates in 11 pivotal RCTs to quantify the potential gain in statistical power from using RAM as the primary outcome. To this end, we employed two approaches: an aggregated-data (AD) approach that combined population-level data from RCT with observational data from the OFSEP registry, and an individual patient data (IPD) approach for a subset of trials.

## METHODS

### RCTs selection

We analyzed 11 pivotal RCTs evaluating five distinct DMTs in relapsing-remitting MS (RRMS): natalizumab (vs. placebo [AFFIRM^2^]); cladribine (vs. placebo [CLARITY^4^]); dimethyl fumarate (DMF) (vs. placebo [DEFINE^6^, CONFIRM^6^] and vs. glatiramer acetate [CONFIRM^6^]); teriflunomide (vs. placebo [TEMSO^5^, TOWER^8^] and vs. interferon [TENERE^7^]), fingolimod (vs. placebo [FREEDOMS^9^], vs. glatiramer acetate [ASSESS^11^], and vs. interferon [TRANSFORMS^3^]); and ocrelizumab (vs. interferon [OPERA^10^]). Trials were selected if DMTs they evaluated were sufficiently represented in the OFSEP ancillary study or if IPD were available in the PRIMUS database. The PRIMUS (PRojections In Multiple Sclerosis) project is a French precision-medicine platform that integrates IPD from multiple RCTs and observational data.^16,17^ Four RCTs were eligible in the PRIMUS database: three (AFFIRM, CONFIRM, and DEFINE) were analyzed using both AD and IPD and one (CLARITY) only using IPD.

### Aggregated-data approach

#### Rationale and overview of the AD approach

The AD approach was designed to estimate RAM and ACES rates in each RCT arm assuming that each relapse had been explored with brain and spinal cord MRI. In a measurement error framework, clinically defined relapse is an indirect measure of RAM, the *true* outcome, with a perfect sensitivity (no events are missed) but imperfect specificity (a proportion of clinically defined relapses are misclassified, corresponding to ACES instead of RAM).^18^ Furthermore, its specificity is differential, meaning that it varies with patient characteristics (age, disability, and disease duration) and, most critically, with DMT.^14^ The AD approach aimed to estimate the expected proportion of RAM and ACES events among clinically defined relapses according to population characteristics and DMT.

The first step was to construct a RAM model, *i.e.* a model of the probability that a clinical event would be classified as RAM rather than ACES, as a function of patient characteristics. We built this model using observational data from the OFSEP registry, which served as an ancillary study in which the actual RAM/ACES classification of clinically defined relapses was known. The RAM model was then employed in each RCT arm using population-level patient characteristics to predict the expected proportion of RAM. Finally, the reported clinically defined relapse rate was multiplied by the expected RAM probability to estimate RAM and ACES rates in each trial arm, which were compared to estimate the treatment effect on RAM and ACES rates.

#### Data source

For each RCT, we extracted aggregated baseline characteristics (age, disease duration, Expanded Disability Status Scale [EDSS]) and relapse rates from published manuscripts. The ancillary study used the same dataset as our previously published study^14^ from the OFSEP database extracted on June 8, 2023.^19^ We included all clinical event reported as a relapse, occurring between January 1, 2015, and June 8, 2023, in patients with a relapsing-remitting MS, with a reference brain MRI within 12 months and spinal cord MRI within 24 months before the event, and a brain and spinal cord MRI with gadolinium injection within 50 days after. Events were excluded if reported as optic neuritis or if ≥2 clinical events occurred between pre– and post-event MRIs. Index clinical events were classified as RAM if the post-event brain or spinal cord MRI indicated worsening, a new or enlarged T2-lesion, and/or a gadolinium-enhanced T1-lesion compared to the pre-event MRI, or as ACES otherwise.

#### RAM model construction

The RAM model was a logistic regression of the probability that a clinical event would be classified as RAM vs. ACES, adjusted for DMT, age, disease duration, and EDSS before the index event. A generalized estimating equations (GEE) approach with an unstructured working matrix was used to account for multiple events occurring in the same patient. Missing EDSS values were imputed using a random forest algorithm to predict EDSS as a function of age, sex, disease duration, DMT, and brain and spinal cord MRI lesion-load.

#### Prediction of the expected RAM probability

For each RCT arm, the expected probabilities of RAM and ACES were predicted using the RAM model. We simulated a population with age, disease duration, and EDSS drawn from a multinormal distribution with the means/standard-deviations of the RCT population, and with a covariance matrix derived from an observational population from the OFSEP database that fulfilled the same inclusion criteria. Then, using the RAM model, a probability of RAM was predicted for each simulated patient to obtain an expected distribution of RAM probability. Finally, a Monte Carlo procedure was applied by randomly drawing a large number of values of RAM probability (from the distribution obtained in the simulated population) and clinically defined relapse rate (from a normal distribution derived from RCT-reported values). This yielded expected RAM and ACES rates for each arm, as well as relative rates and 95% confidence intervals (CI).

### Individual-patient data approach

#### Data sources

The PRIMUS platform integrates multiple trial datasets using a harmonizing pipeline, ensuring consistent variable definitions and formats. For each patient in the four available RCTs, we used data on relapse occurrence, brain MRI (new/enlarged T2-lesions and new gadolinium-enhanced T1-lesions), and treatment arm. Each clinically defined relapse was classified as RAM if radiological activity was detected on the subsequent brain MRI, as ACES if no radiological activity was reported, or as unclassified if no post-event MRI was available.

#### RAM and ACES rates

For each trial arm, to obtain RAM and ACES rates despite unclassified events, the expected probability of RAM was derived from the proportion of events classified as RAM vs. ACES. A Monte Carlo procedure was used to obtain the expected RAM and ACES rates and their 95% CIs by drawing a large sample of RAM probability and relapse rate values, assuming a normal distribution. Relative RAM and ACES rate between trial arms were estimated directly from classified events using an univariate Poisson regression.

### Statistical power analysis

To assess the statistical power gain from using RAM as the primary outcome instead of clinically defined relapse, we compared theoretical sample sizes and trial duration requirements across different scenarios of ARR in the treated and control arm. We set a statistical power of 90% with 5% two-sided alpha risk, including 20% loss to follow-up, and assuming equal ACES rates of 0.10 events/year in both arms and a Poisson distribution for relapses.

### Statistical analysis

P-values below 0.05 were considered statistically significant. Analyses were performed using R software, version 4.4.2,^20^ with *geepack*^21^ and *rpact*^22^ packages.

## RESULTS

### AD approach

From the OFSEP database, 637 clinical events from 608 patients were included to fit the RAM model, of which 471 (73.9%) were classified as RAM. Mean age was 35.8 (SD, 10.7) years, mean disease duration was 5.6 (6.6) years, and mean EDSS score was 1.9 (1.6) (eTable 1 in Supplement). Regarding DMT exposure, 273 (42.9%) events occurred in untreated patients, 43 (6.8%) were receiving interferon, 70 (11.0%) glatiramer acetate, 63 (9.9%) teriflunomide, 54 (8.5%) DMF, 59 (9.3%) fingolimod, 32 (5.0%) natalizumab, and 24 (3.8%) intravenous anti-CD20. We imputed 181 (28.4%) missing EDSS measurements. The details of the RAM logistic model are provided in eTable 2 in the Supplement.

### IPD approach

Among 3675 patients included in the four trials, 2018 clinically defined relapses were analyzed: 864 (42.8%) were classified as RAM, 513 (25.4%) as ACES, and 641 (31.8%) were unclassified (Table 1). Unclassified events occurred mainly in the CONFIRM and DEFINE trials, where only a subset had MRI follow-up. The median interval between relapse and post-event MRI was 144 days (IQR, 73-250) and was similar for both RAM and ACES. Events were significantly more likely to be classified as ACES in older patients (Wilcoxon test, p < 0.001), with longer disease duration (p < 0.001), and higher EDSS score before the event (p < 0.001).

**Table 1.**
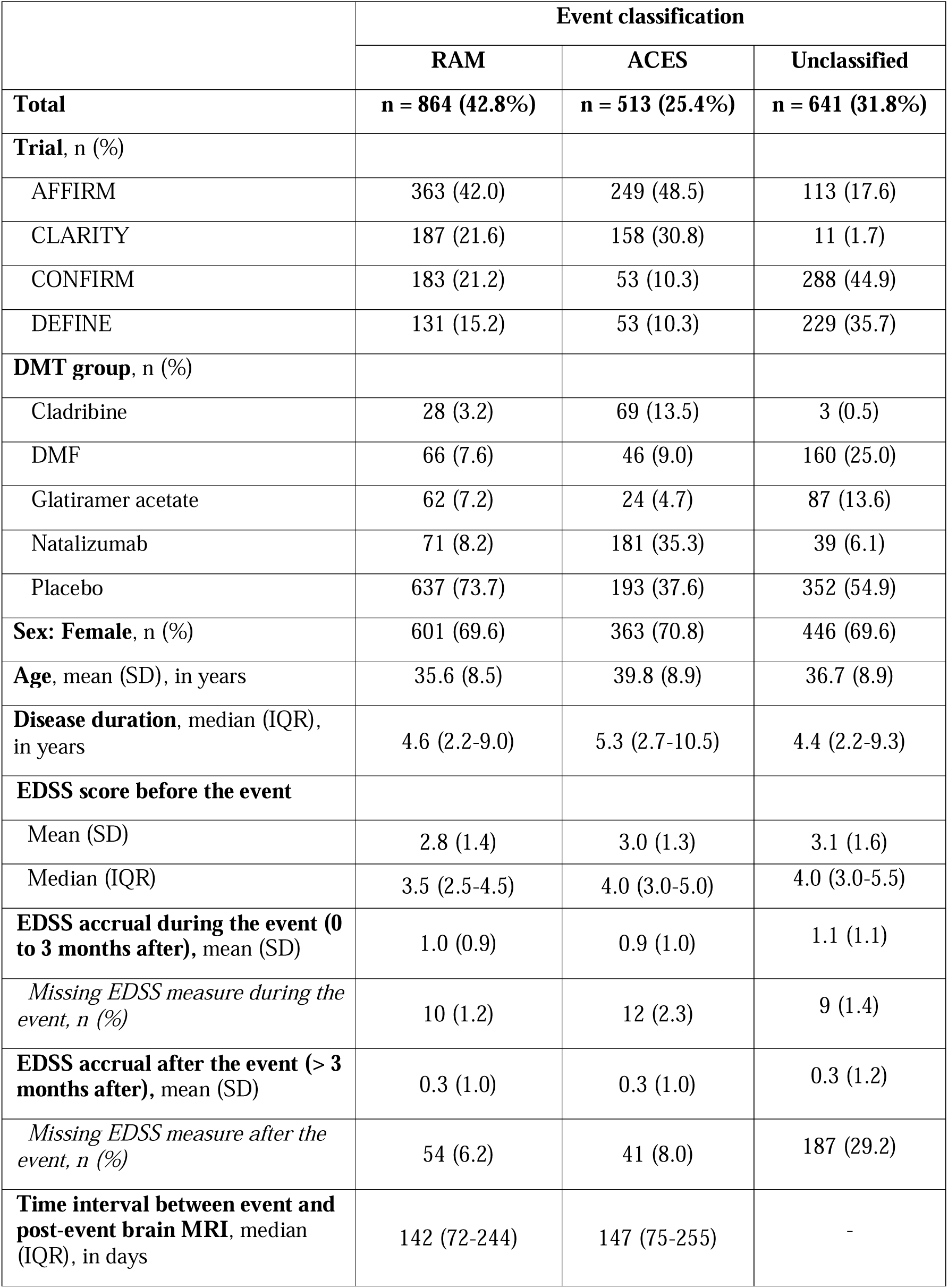
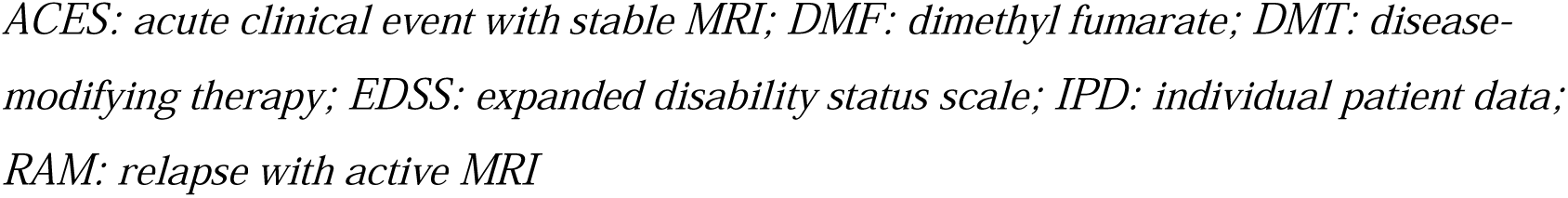
Description of clinically defined relapses according to their classification in the IPD analysis of the AFFIRM, CLARITY, CONFIRM, and DEFINE trials.

|  | Event classification |  |  |
| --- | --- | --- | --- |
|  | RAM | ACES | Unclassified |
| **Total** | **n = 864 (42.8%)** | **n = 513 (25.4%)** | **n = 641 (31.8%)** |
| **Trial, n (%)** |  |  |  |
| AFFIRM | 363 (42.0) | 249 (48.5) | 113 (17.6) |
| CLARITY | 187 (21.6) | 158 (30.8) | 11 (1.7) |
| CONFIRM | 183 (21.2) | 53 (10.3) | 288 (44.9) |
| DEFINE | 131 (15.2) | 53 (10.3) | 229 (35.7) |
| **DMT group, n (%)** |  |  |  |
| Cladribine | 28 (3.2) | 69 (13.5) | 3 (0.5) |
| DMF | 66 (7.6) | 46 (9.0) | 160 (25.0) |
| Glatiramer acetate | 62 (7.2) | 24 (4.7) | 87 (13.6) |
| Natalizumab | 71 (8.2) | 181 (35.3) | 39 (6.1) |
| Placebo | 637 (73.7) | 193 (37.6) | 352 (54.9) |
| **Sex: Female, n (%)** | 601 (69.6) | 363 (70.8) | 446 (69.6) |
| **Age, mean (SD), in years** | 35.6 (8.5) | 39.8 (8.9) | 36.7 (8.9) |
| **Disease duration, median (IQR), in years** | 4.6 (2.2-9.0) | 5.3 (2.7-10.5) | 4.4 (2.2-9.3) |
| **EDSS score before the event** |  |  |  |
| Mean (SD) | 2.8 (1.4) | 3.0 (1.3) | 3.1 (1.6) |
| Median (IQR) | 3.5 (2.5-4.5) | 4.0 (3.0-5.0) | 4.0 (3.0-5.5) |
| **EDSS accrual during the event (0 to 3 months after), mean (SD)** | 1.0 (0.9) | 0.9 (1.0) | 1.1 (1.1) |
| *Missing EDSS measure during the event, n (%)* | 10 (1.2) | 12 (2.3) | 9 (1.4) |
| **EDSS accrual after the event (> 3 months after), mean (SD)** | 0.3 (1.0) | 0.3 (1.0) | 0.3 (1.2) |
| *Missing EDSS measure after the event, n (%)* | 54 (6.2) | 41 (8.0) | 187 (29.2) |
| **Time interval between event and post-event brain MRI, median (IQR), in days** | 142 (72-244) | 147 (75-255) | - |

*ACES: acute clinical event with stable MRI; DMF: dimethyl fumarate; DMT: disease-modifying therapy; EDSS: expanded disability status scale; IPD: individual patient data; RAM: relapse with active MRI*

### Expected RAM probabilities

The expected probability for a clinical relapse to be classified as RAM estimated by the AD approach varied considerably across RCT arms (eTable 3 in Supplement). It was lowest in arms receiving a highly effective DMT: only 25.3% in the ocrelizumab arm of the OPERA trial, and approximately 45% in arms receiving natalizumab (AFFIRM) or fingolimod (ASSESS, FREEDOMS, TRANSFORM). In contrast, arms receiving lower-efficacy DMTs or placebo had a higher expected probability of RAM: approximately 70% for teriflunomide, 75% for interferon, 80% for glatiramer acetate and DMF, and 82% for placebo arms. With the IPD approach in the AFFIRM, CONFIRM, and DEFINE trials, expected RAM probabilities estimated were lower for treated arms than with the AD approach (by 15 to 30 points): 28.2% in the natalizumab arm, 67.9% in the DMF arm of the CONFIRM trial, and 50.8% in the DMF arm of the DEFINE trial.

### Treatment effect on RAM and ACES rates

Treatment effects had a similar direction and statistical significance on clinically defined relapse and on RAM (Table 2). Furthermore, in 7 of the 10 RCTs, the magnitude of the treatment effect was greater on RAM. This reached up to an almost threefold increase in effect size in the OPERA trial, with a relative rate (RR) of 0.20 (95% CI, 0.07-0.42) for RAM compared with 0.54 (95% CI, 0.43-0.66) for clinically defined relapses. For the 3 remaining trials, treatment effects were similar on RAM or clinically defined relapses: in the CONFIRM and DEFINE trials, both of which evaluated DMF, and in the TENERE trial, whose results were inconclusive. With the IPD approach, treatment effects on RAM rate were larger in magnitude than those on clinically defined relapses, including the effect of DMF in the CONFIRM and DEFINE trials, which reached statistical significance in the comparison of DMF vs. glatiramer acetate. IPD-estimated effects were also of slightly larger magnitude than those estimated with the AD approach.

**Table 2.**
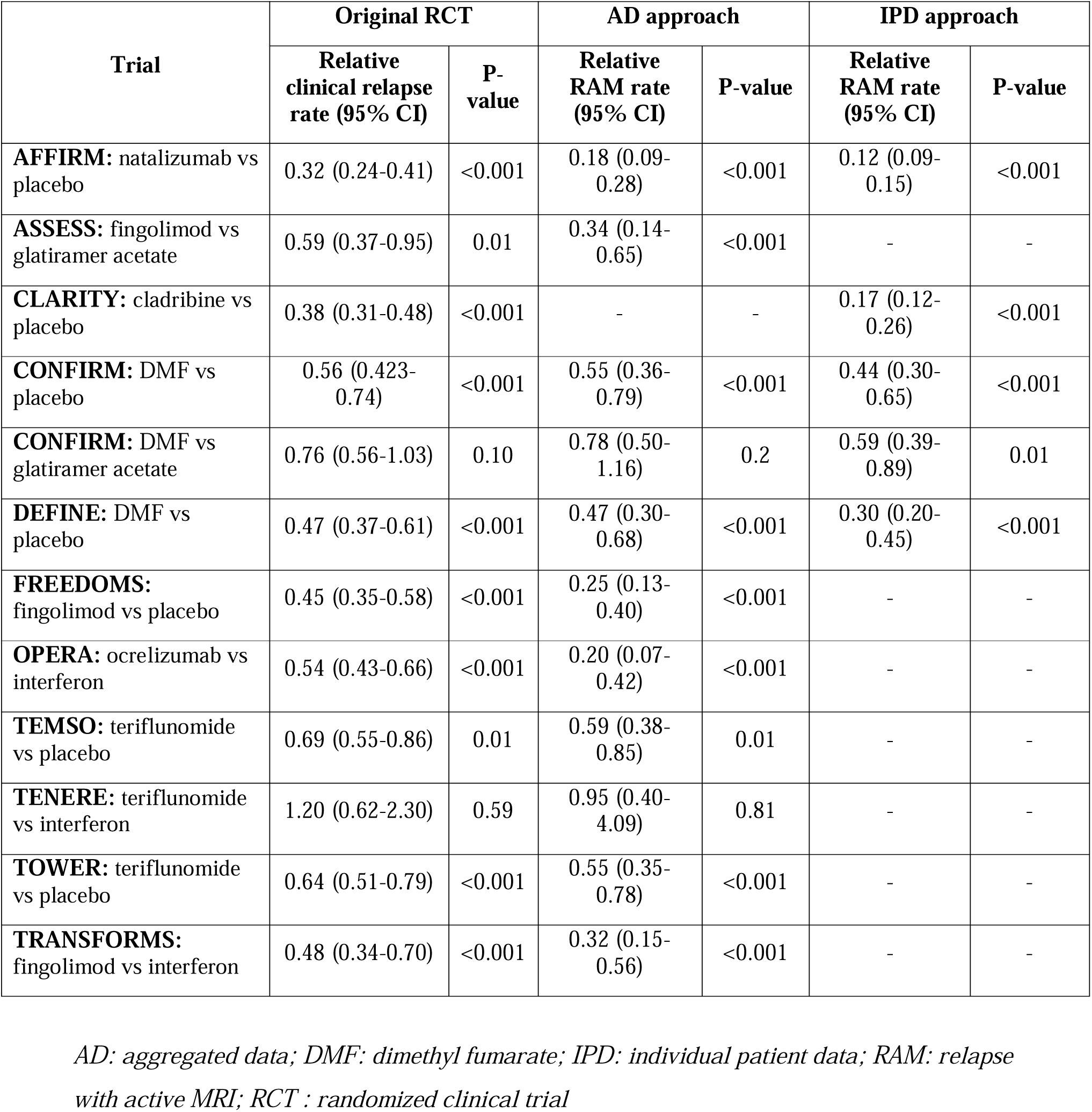
Estimated treatment effect on clinically defined annualized relapse rate and RAM rate in each trial.

Conversely, expected ACES rates were similar across all RCT arms. The average ACES rate estimated with the AD approach was 0.08 event/year, ranging from 0.04 event/year in the DMF arm of the DEFINE trial to 0.13 event/year in the placebo arm of the AFFIRM trial (Figure 1). No DMT were found to have a statistically significant effect on ACES rate (eTable 3 in Supplement). Results were consistent with the IPD approach, although expected ACES rates were slightly higher, with an average of 0.09 event/year.

**Figure 1.**
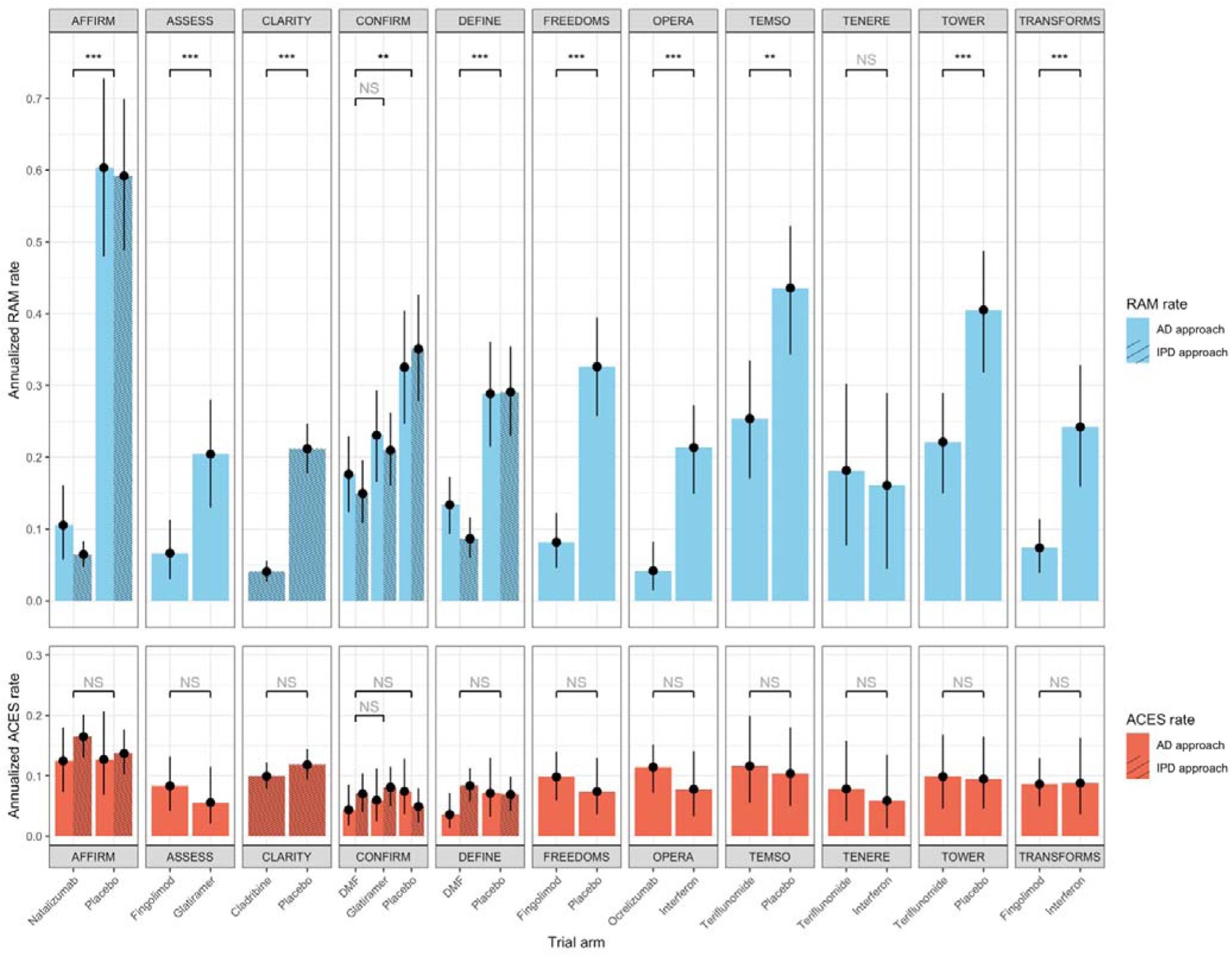
Estimated annualized RAM and ACES rates in each trial. *** represents p-value < 0.001, ** p-values < 0.01, * p-values < 0.05, and NS: not significant For trials evaluated using both the AD and IPD approaches, the results of statistical comparison were similar. *ACES: acute clinical event with stable MRI; AD: aggregated data; DMF: dimethyl fumarate; IPD: individual patient data; RAM: relapse with active MRI*

### Statistical power gain using RAM as the primary outcome

Using RAM as the primary outcome increased statistical power to detect a treatment effect compared to clinically defined relapses across most scenarios of ARR in the treated and control arms (eFigure 1 in Supplement). This power gain increased as the ARR in the control arm decreased (Figure 2). The largest sample size reduction reached 55.7% in the scenario with an ARR of 0.15 in the treated arm compared to 0.20 in the control arm. Notably, based on ARRs observed in the 10 conclusive RCTs, using RAM as the primary outcome would have enabled a theoretical reduction in the required sample size by 30% to 45% for the ASSESS, CLARITY, DEFINE, OPERA, and TRANSFORMS trials, by 20% to 30% for the CONFIRM, FREEDOMS, TEMSO, and TOWER trials, and by approximately 13% for the AFFIRM trial. For instance, in a scenario analogous to the OPERA trial (ARR of 0.15/year in the treated arm vs. 0.30/year in the control arm), using clinically defined relapse as the primary outcome would require 440 patients followed for 2 years to obtain a 90% power, while using RAM instead would require only 276 patients followed for 2 years or the same number of patients for 1.25 year (Figure 3).

**Figure 2.**
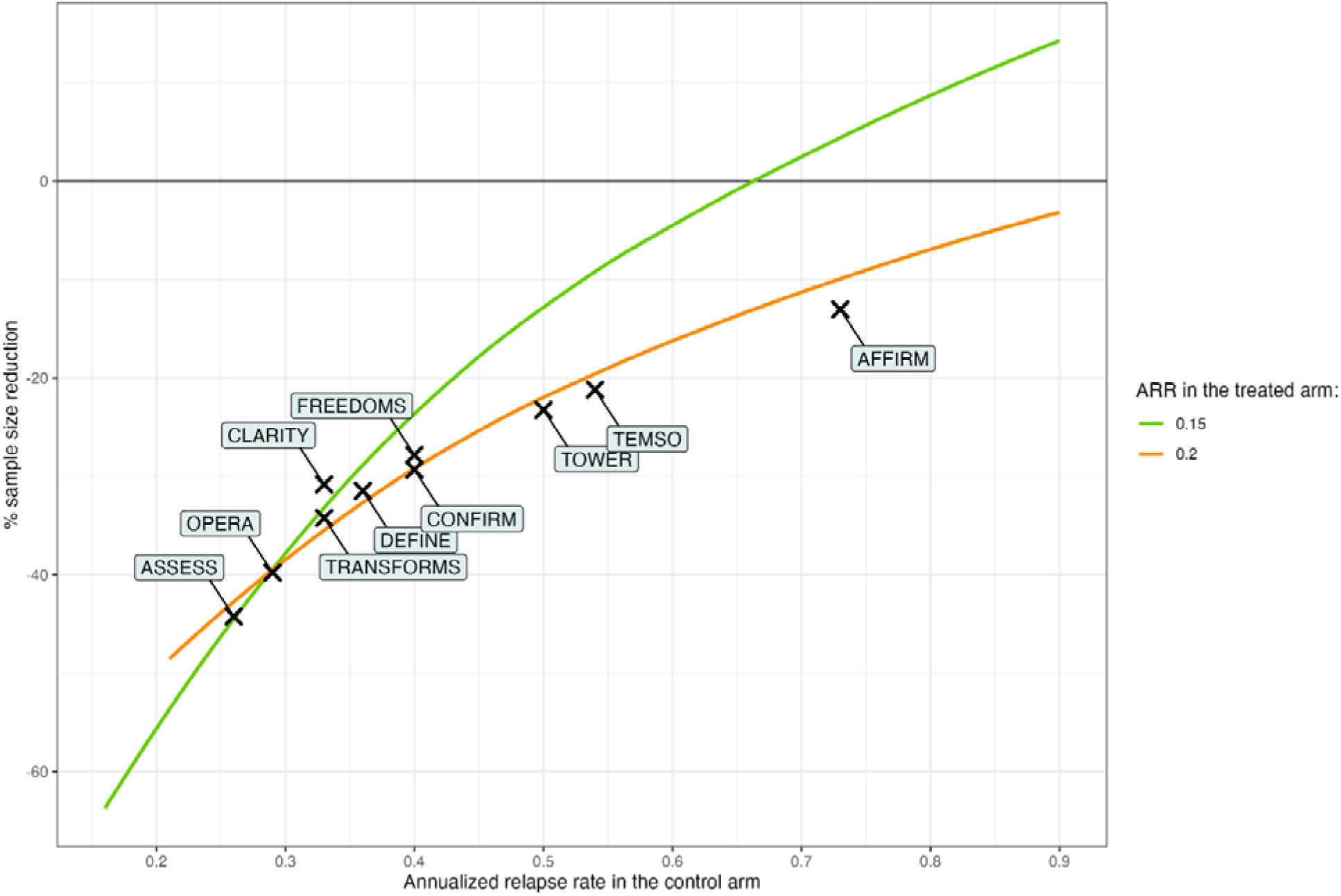
Sample size reduction with RAM as the main outcome compared to clinically defined relapses according to the ARR in the control arm. To achieve a statistical power of 90%, with a 5% two-sided alpha risk, with an ACES rate of 0.10/year in each arm, and for two ARR values in the treated arm of 0.15 and 0.20/year. *ACES: acute clinical event with stable MRI; ARR: annualized relapse rate, RAM: relapse with active MRI*

**Figure 3.**
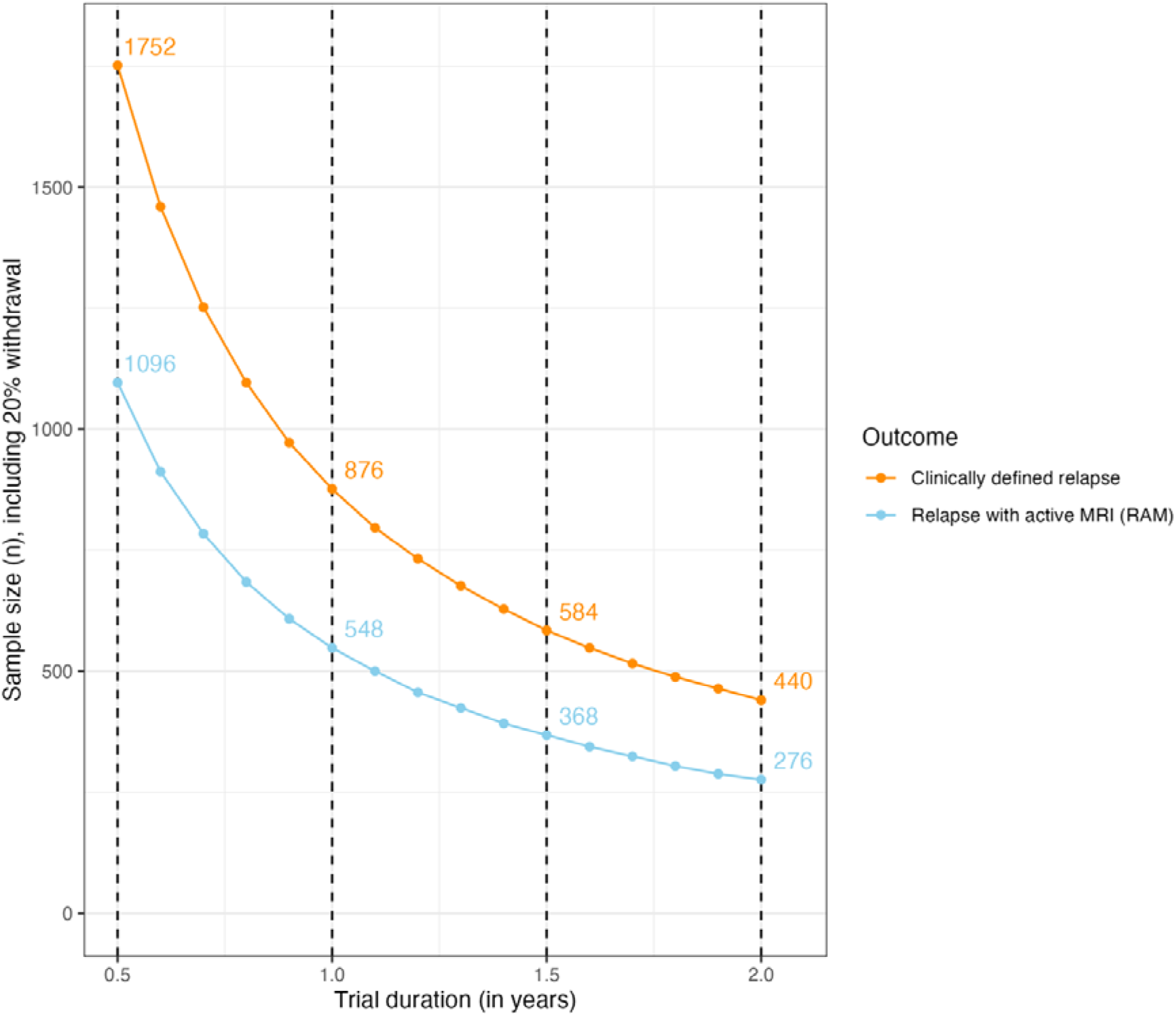
Calculation of the sample size and trial duration required to achieve a 90% power with clinically defined relapses or RAM as the primary outcome. To achieve a statistical power of 90%, with a 5% two-sided alpha risk, annualized relapse rates of 0.15/year in the active arm and 0.30/year in the control arm, ACES rates of 0.10/year in each arm (equivalent to RAM rates of 0.05/year in the active arm and 0.20/year in the control arm), and including 20% of loss to follow-up. *ACES: acute clinical event with stable MRI; ARR: annualized relapse rate, RAM: relapse with active MRI; RR: relative rate*

## DISCUSSION

In this study, we estimated RAM and ACES rates in each arm of 11 RCTs by applying two distinct approaches based on AD or IPD. Our findings indicate that treatment effects on RAM were equivalent to or greater than those on clinically defined relapses. In contrast, no DMT affected the ACES rate, stable at ∼0.08/year across all arms. Employing RAM as the primary outcome could enhance the statistical power of future RCTs in MS, theoretically reducing the required sample size or trial duration by 30 to 50% in scenarios reflecting ARR observed in recent trials.

In accordance with our previous study,^14^ our results suggest that RAM is the primary target of current DMTs, while ACES act as background noise that dilutes treatment effect estimates. By incorporating ACES within its definition, clinically defined relapses might underestimate treatment effects on inflammatory activity, particularly in recent MS trials, where lower-than-expected relapse rates in the control arm have led to inconclusive results for promising new therapies.^12^ Several factors may explain this trend, including the use of active comparators, selection of patients with less active disease (as patients with active disease may prefer available, highly effective DMTs over participating in a trial), and changes in MS diagnostic criteria over time. Given these challenges, systematically evaluating each clinical event with a comprehensive brain and spinal cord MRI to assess RAM as the primary outcome could provide a methodological advantage for future trials, supporting further DMT development to offer patients a wider choice of therapeutic options.

Primary endpoints relying solely on MRI activity, mainly new/enlarging T2-lesions and gadolinium-enhanced T1-lesions, are common in phase 2 trials. Prior research has shown that MRI-based outcomes could serve as surrogate markers for relapses.^23,24^ However, they primarily capture subclinical disease activity, which may not always translate into functional impairment for patients. By integrating clinical and radiological components, RAM events are both biologically and clinically relevant. Moreover, MRIs triggered by a clinical event are more likely than scheduled MRIs to detect radiological activity, particularly gadolinium-enhanced lesions or detection guided by the patient’s symptoms.

The present study provided the opportunity to compare two distinct approaches for evaluating ACES and RAM events in trials. The AD approach relied on OFSEP observational data with strict selection criteria for MRI timing and completeness (mainly spinal cord assessment and gadolinium injection) in order to maximize sensitivity for detecting radiological activity. In contrast, the IPD approach took advantage of the strict MRI monitoring in RCTs to analyze a less selected population, in which the imaging decision was independent of the clinical event characteristics. However, no spinal cord MRIs were performed, and brain MRIs were obtained only at predefined intervals, generally annually or semiannually, thus reducing the sensitivity for detecting radiological activity. As expected, these differences resulted in lower RAM probabilities estimated using the IPD approach than those estimated using the AD approach, and slightly higher ACES rates. Additionally, the IPD approach may be beneficial for clarifying results of recent trials of emerging therapies, such as Bruton’s tyrosine kinase inhibitor, by distinguishing their effects on inflammatory and non-inflammatory components of the disease.^12,13^

Our study has several limitations. First, the RAM model was developed using observational data, introducing a risk of confounding bias between DMT groups, which could propagate to RAM and ACES estimates in RCTs. Additionally, several factors could have limited the transportability of the RAM model to the RCT setting, such as unmeasured covariates that could alter ACES/RAM probabilities, exclusion of optic neuritis, and differences in the diagnostis of relapses between clinical practice and RCTs. In particular, the AD approach estimated higher RAM probabilities under DMF than with other DMTs, resulting in larger differences in ACES rates. However, the IPD analysis did not confirmed these findings.

Second, the AD approach used baseline characteristics of the entire RCT population, which may not reflect the distribution of age, disease duration, and EDSS of patients at the time of relapse. However, in our model, DMT was the main determinant of the RAM probability, whereas EDSS score, age, and disease duration had only a minor influence. Consequently, our study should be seen as a preliminary exploration of shifting the primary outcome from clinically defined relapses to MRI-confirmed relapses in MS trials, and larger studies are warranted to confirm or refute our findings.

In conclusion, adopting RAM instead of clinically defined relapses as the primary outcome could be a valuable methodological choice for future RCTs in MS, increasing the statistical power to detect treatment effects. Moreover, incorporating a systematic brain and spinal cord MRI assessment following each clinical event could provide crucial insights for secondary analyses, particularly in defining actual DMT targets and clarifying their mechanisms of action.

## Data Availability

Data may be made available at the motivated request by any expert researcher, after approval by the OFSEP and PRIMUS.

## Acknowledgments

Data collection has been supported by a grant provided by the French State and handled by the *Agence Nationale de la Recherche*, within the framework of the *France 2030* program, under the reference ANR-10-COHO-002, *Observatoire Français de la Sclérose en Plaques* (OFSEP) and by the Eugène Devic EDMUS Foundation against multiple sclerosis.

## Data sharing statement

Data may be made available at the motivated request of any expert researcher, after approval by the OFSEP and the PRIMUS committees.

## Contributions

AG, MN and DL designed the study; all authors collected the data; AG performed the statistical analysis; AG and DL interpreted the data; AG and DL drafted the manuscript; all authors reviewed the manuscript; AG and DL had full access to all the data and take responsibility for the integrity of the data and the accuracy of the data analysis.

## Disclosures

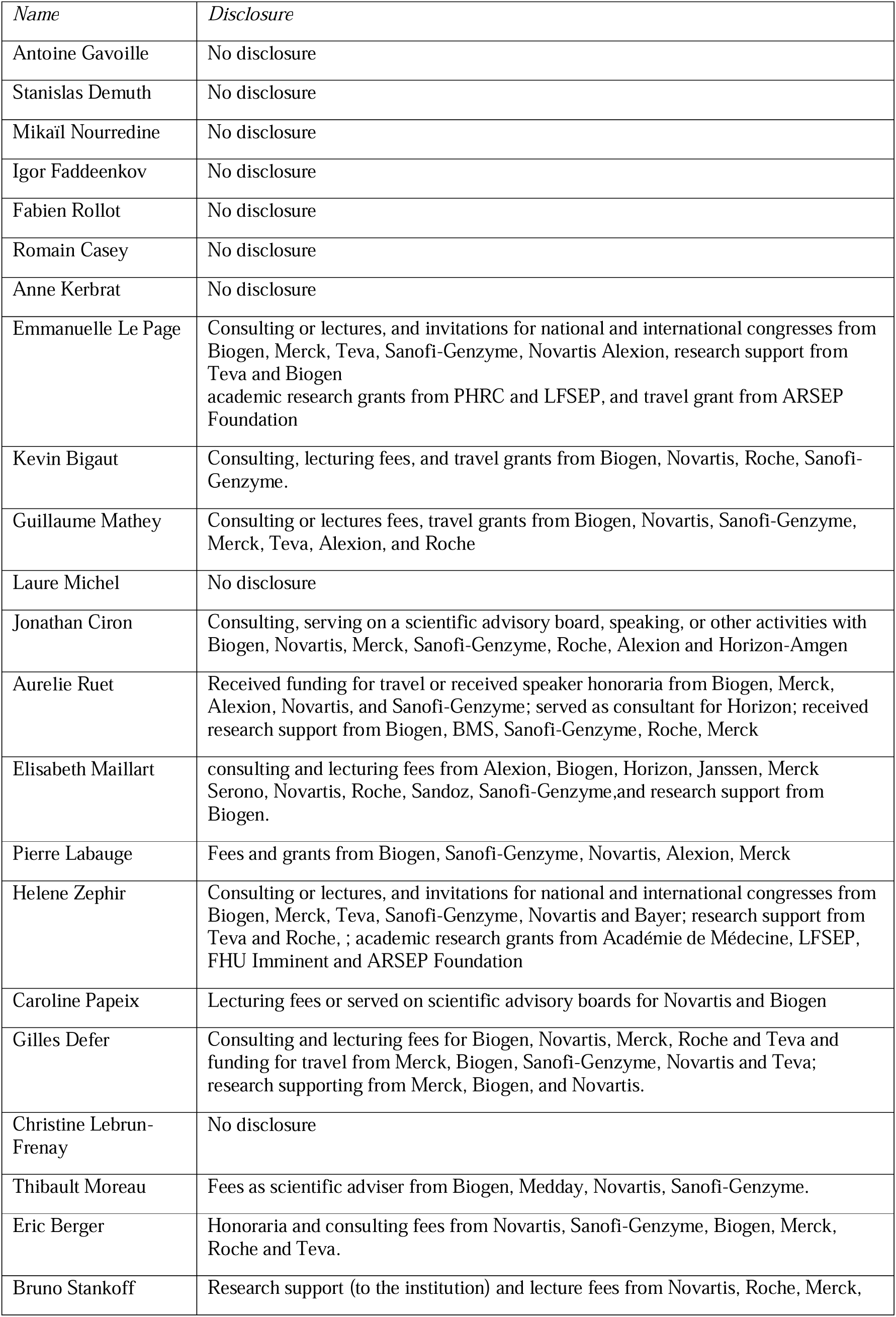
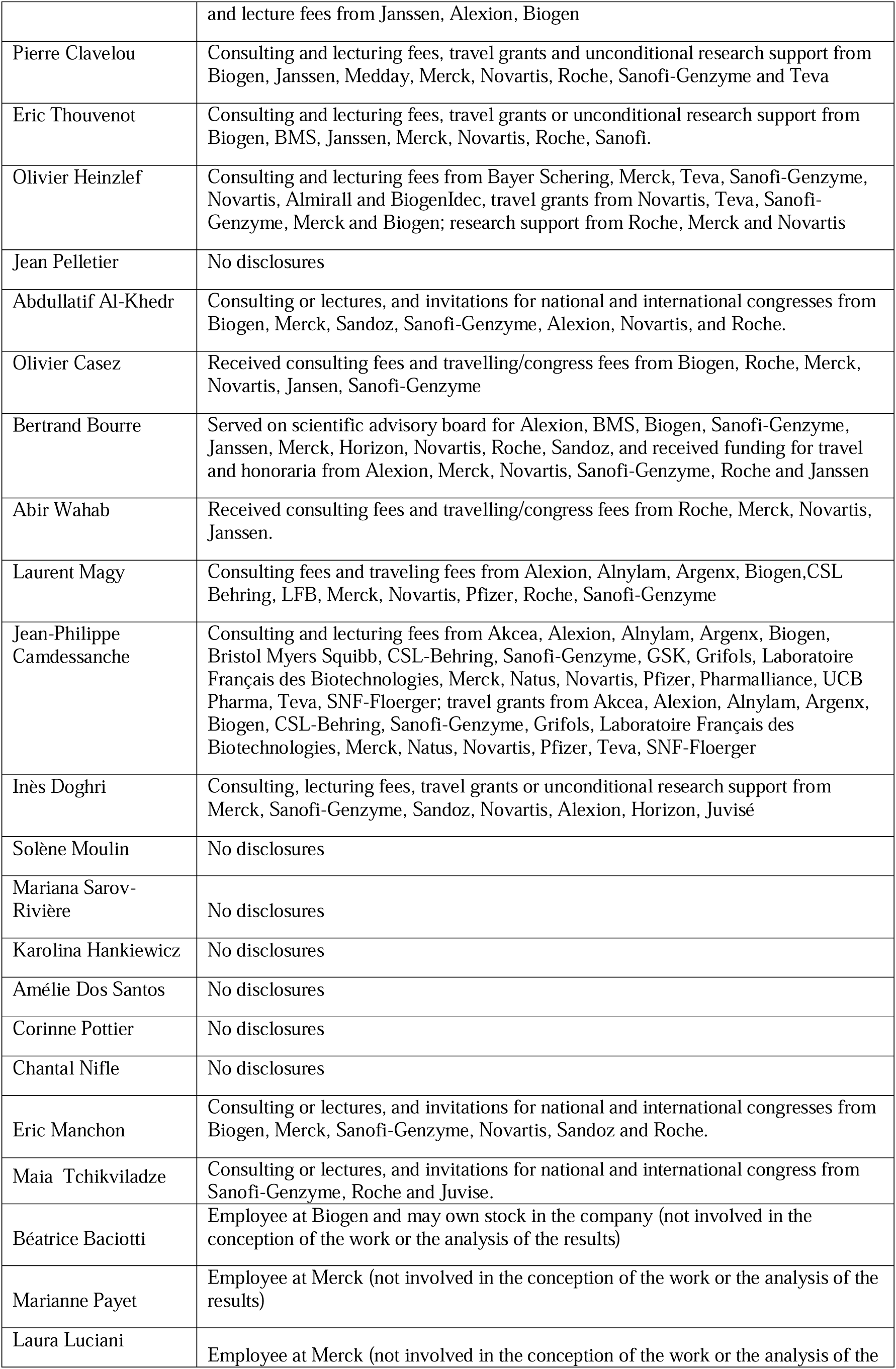
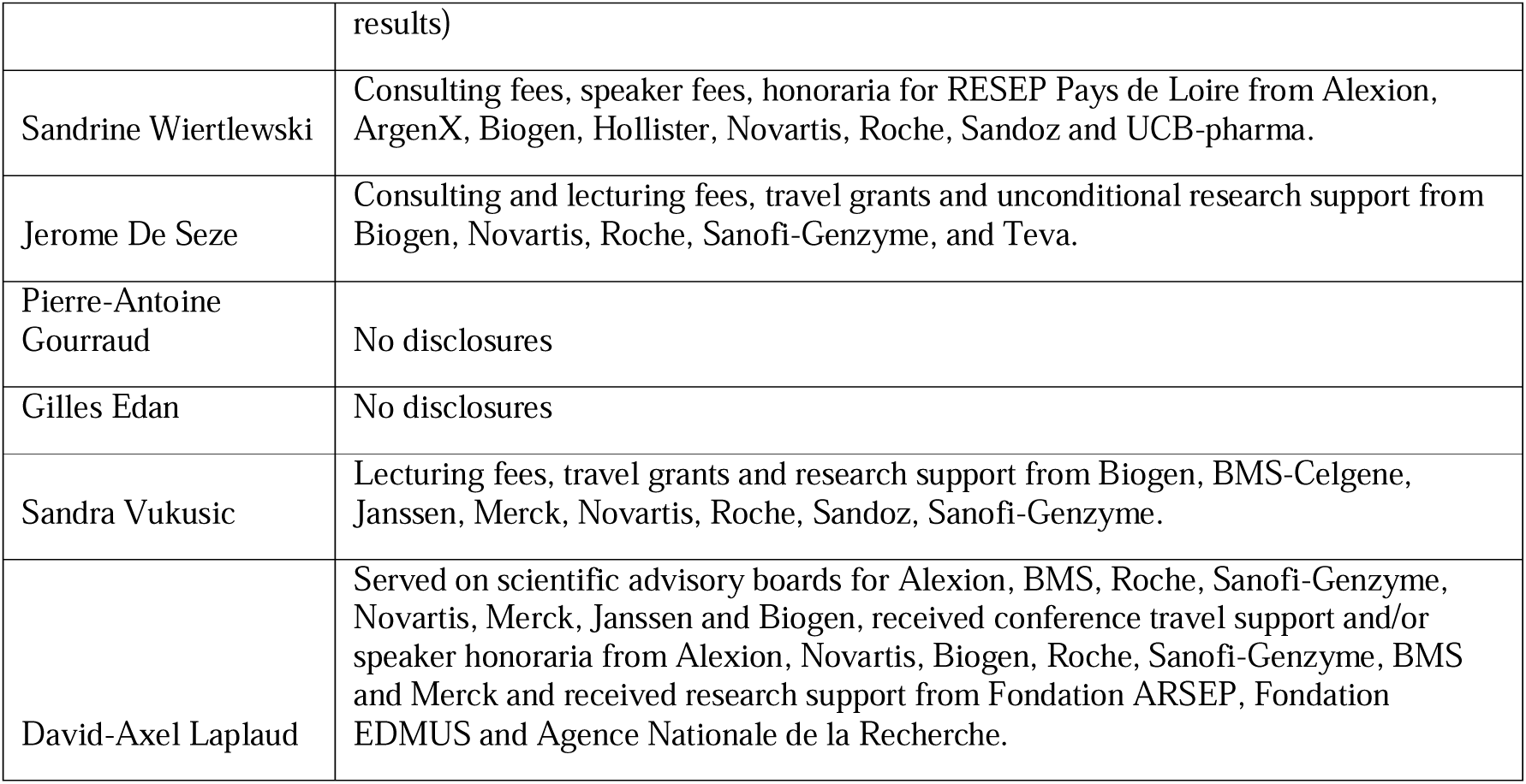

## REFERENCES

1. Thompson AJ, Banwell BL, Barkhof F, et al. Diagnosis of multiple sclerosis: 2017 revisions of the McDonald criteria. Lancet Neurol. 2018;17(2):162–173. doi:10.1016/S1474-4422(17)30470-2

2. Polman CH, O’Connor PW, Havrdova E, et al. A Randomized, Placebo-Controlled Trial of Natalizumab for Relapsing Multiple Sclerosis. N Engl J Med. 2006;354(9):899–910. doi:10.1056/NEJMoa044397

3. Cohen JA, Barkhof F, Comi G, et al. Oral Fingolimod or Intramuscular Interferon for Relapsing Multiple Sclerosis. N Engl J Med. 2010;362(5):402–415. doi:10.1056/NEJMoa0907839

4. Giovannoni G, Comi G, Cook S, et al. A Placebo-Controlled Trial of Oral Cladribine for Relapsing Multiple Sclerosis. N Engl J Med. 2010;362(5):416–426. doi:10.1056/NEJMoa0902533

5. O’Connor P, Comi G, Benzerdjeb H, Miller A. Randomized Trial of Oral Teriflunomide for Relapsing Multiple Sclerosis. N Engl J Med. Published online 2011.

6. Fox RJ, Miller DH, Phillips JT, et al. Placebo-Controlled Phase 3 Study of Oral BG-12 or Glatiramer in Multiple Sclerosis. N Engl J Med. 2012;367(12):1087–1097. doi:10.1056/NEJMoa1206328

7. Vermersch P, Czlonkowska A, Grimaldi LM, et al. Teriflunomide versus subcutaneous interferon beta-1a in patients with relapsing multiple sclerosis: a randomised, controlled phase 3 trial. Mult Scler J. 2014;20(6):705–716. doi:10.1177/1352458513507821

8. Confavreux C, O’Connor P, Comi G, et al. Oral teriflunomide for patients with relapsing multiple sclerosis (TOWER): a randomised, double-blind, placebo-controlled, phase 3 trial. Lancet Neurol. 2014;13(3):247–256. doi:10.1016/S1474-4422(13)70308-9

9. Calabresi PA, Radue EW, Goodin D, et al. Safety and efficacy of fingolimod in patients with relapsing-remitting multiple sclerosis (FREEDOMS II): a double-blind, randomised, placebo-controlled, phase 3 trial. Lancet Neurol. 2014;13(6):545–556. doi:10.1016/S1474-4422(14)70049-3

10. Hauser SL, Bar-Or A, Comi G, et al. Ocrelizumab versus Interferon Beta-1a in Relapsing Multiple Sclerosis. N Engl J Med. 2017;376(3):221–234. doi:10.1056/NEJMoa1601277

11. Cree BAC, Goldman MD, Corboy JR, et al. Efficacy and Safety of 2 Fingolimod Doses vs Glatiramer Acetate for the Treatment of Patients With Relapsing-Remitting Multiple Sclerosis: A Randomized Clinical Trial. JAMA Neurol. 2021;78(1):48. doi:10.1001/jamaneurol.2020.2950

12. Montalban X, Vermersch P, Arnold DL, et al. Safety and efficacy of evobrutinib in relapsing multiple sclerosis (evolutionRMS1 and evolutionRMS2): two multicentre, randomised, double-blind, active-controlled, phase 3 trials. Lancet Neurol. 2024;23(11):1119–1132. doi:10.1016/S1474-4422(24)00328-4

13. Oh J, Arnold DL, Cree BAC, et al. Tolebrutinib versus Teriflunomide in Relapsing Multiple Sclerosis. N Engl J Med. 2025;392(19):1893–1904. doi:10.1056/NEJMoa2415985

14. Gavoille A, Rollot F, Casey R, et al. Acute Clinical Events Identified as Relapses With Stable Magnetic Resonance Imaging in Multiple Sclerosis. JAMA Neurol. 2024;81(8):814. doi:10.1001/jamaneurol.2024.1961

15. Keogh RH, Shaw PA, Gustafson P, et al. STRATOS guidance document on measurement error and misclassification of variables in observational epidemiology: Part 1— Basic theory and simple methods of adjustment. Stat Med. 2020;39(16):2197–2231. doi:10.1002/sim.8532

16. Demuth S, Ed-Driouch C, Rousseau O, et al. PRIMUS-Alpha[: prototype de médecine de précision dans la sclérose en plaques contextualisant l’évolution des patients dans des données de référence multi-sources. Rev Neurol (Paris). 2023;179:S152. doi:10.1016/j.neurol.2023.01.670

17. Demuth S, Faddeenkov I, Paris J, et al. Are we in a Big Data era for multiple sclerosis? Lessons from integrating clinical trials and observational studies data into the PRIMUS precision medicine platform. Preprint posted online October 19, 2024. doi:10.1101/2024.10.17.24315655

18. Nourredine M, Subtil F, Gavoille A, Lepage C, Kassai-Koupai B, Cucherat M. Accounting for Misclassification of Binary Outcomes in External Control Arm Studies for Unanchored Indirect Comparisons: Simulations and Applied Example. Stat Med. Published online 2025.

19. Vukusic S, Casey R, Rollot F, et al. Observatoire Français de la Sclérose en Plaques (OFSEP): A unique multimodal nationwide MS registry in France. Mult Scler J. 2020;26(1):118–122. doi:10.1177/1352458518815602

20. R: A language and environment for statistical computing. R Foundation for Statistical Computing. Published online 2020. https://www.R-project.org/

21. Halekoh U, Højsgaard S, Yan J. The *R* Package **geepack** for Generalized Estimating Equations. J Stat Softw. 2006;15(2). doi:10.18637/jss.v015.i02

22. Wassmer G, Pahlke F. rpact: Confirmatory Adaptive Clinical Trial Design and Analysis. Published online October 30, 2018:4.1.0. doi:10.32614/CRAN.package.rpact

23. Sormani MP, Molyneux PD, Gasperini C, et al. Statistical power of MRI monitored trials in multiple sclerosis: new data and comparison with previous results. J Neurol Neurosurg Psychiatry. 1999;66(4):465–469. doi:10.1136/jnnp.66.4.465

24. Pia Sormani M, Bruzzi P, Comi G, Filippi M. MRI metrics as surrogate markers for clinical relapse rate in relapsing-remitting MS patients. Neurology. 2002;58(3):417–421. doi:10.1212/WNL.58.3.417

