## Supplement for "Evaluating MRI-Confirmed Relapses as a Novel Primary Endpoint in Multiple Sclerosis Trials"

**Supplementary Tables :**

**eTable 1.** Description of patient characteristics at the time of the index event in the OFSEP ancillary study

**eTable 2.** Details of the RAM vs ACES logistic model (on the logit scale)

**eTable 3.** Annualized relapse rates and expected RAM probabilities in RCT populations

**eTable 4.** Estimated treatment effect on ACES rate

**Supplementary Figure :**

**eFigure 1.** Sample size reduction with RAM as the primary outcome compared to clinically defined relapses, with varying ARR in the treated and control arms

### SUPPLEMENTARY TABLES

#### eTable 1. Description of patient characteristics at the time of the index event in the OFSEP ancillary study

| | **RAM** (n = 471) | **ACES** (n = 166) | **Total** (n = 637) |
|---|---|---|---|
| **Age** (in years), mean (SD) | 34.9 (10.5) | 38.2 (10.9) | 35.8 (10.7) |
| **Disease duration** (in years), mean (SD) | 4.8 (5.9) | 7.7 (7.8) | 5.6 (6.6) |
| **EDSS score,** mean (SD) | 1.7 (1.5) | 2.3 (1.7) | 1.9 (1.6) |
| **Missing EDSS score,** n (%) | 147 (31.2) | 34 (20.5) | 181 (28.4) |
| **Imputed EDSS score**, mean (SD) | 1.6 (1.4) | 2.1 (1.7) | 1.7 (1.5) |
| **DMT**, n (%) | | | |
| None | 229 (48.6) | 44 (26.5) | 273 (42.9) |
| Interferon | 33 (7.0) | 10 (6.0) | 43 (6.8) |
| Glatiramer acetate | 58 (12.3) | 12 (7.2) | 70 (11.0) |
| Teriflunomide | 44 (9.3) | 19 (11.4) | 63 (9.9) |
| DMF | 45 (9.6) | 9 (5.4) | 54 (8.5) |
| Fingolimod | 26 (5.5) | 33 (19.9) | 59 (9.3) |
| Natalizumab | 14 (3.0) | 18 (10.8) | 32 (5.0) |
| Intravenous anti-CD20 | 8 (1.7) | 16 (9.6) | 24 (3.8) |
| Other | 14 (3.0) | 5 (3.0) | 19 (3.0) |

*ACES: acute clinical event with stable MRI; DMF: dimethyl fumarate; DMT: disease-modifying therapy; EDSS: expanded disability status scale; RAM: relapse with active MRI*

**eTable 2. Details of the RAM vs ACES logistic model (on the logit scale)**

| | **β (95% CI)** | **P-value** |
|---|---|---|
| **Intercept** | 2.44 (1.68; 3.20) | **-** |
| **Age**, per 10 years | -0.03 (-0.18; 0.12) | 0.69 |
| **Disease duration**, per 10 years | -0.26 (-0.57; 0.05) | 0.10 |
| **EDSS**, per 1 point | -0.17 (-0.38; 0.04) | 0.11 |
| **DMT** | | |
| **None** | Reference | - |
| **Interferon** | -0.48 (-1.28; 0.31) | 0.23 |
| **Glatiramer acetate** | -0.11 (-0.82; 0.6) | 0.76 |
| **DMF** | -0.06 (-0.86; 0.74) | 0.89 |
| **Teriflunomide** | -0.67 (-1.32; -0.01) | 0.04 |
| **Fingolimod** | -1.72 (-2.35; -1.1) | <0.001 |
| **Natalizumab** | -1.77 (-2.55; -0.99) | <0.001 |
| **Intravenous anti-CD20** | -2.64 (-3.69; -1.58) | <0.001 |
| **Other** | -0.58 (-1.67; 0.50) | 0.29 |

*ACES: acute clinical event with stable MRI; DMF: dimethyl fumarate; DMT: disease-modifying therapy; EDSS: expanded disability status scale; RAM: relapse with active MRI*

**eTable 3. Annualized relapse rates and expected RAM probabilities in RCT populations**

| Trial | Annualized relapse rate | | Expected RAM probability (AD approach) | | Expected RAM probability (IPD approach) | |
|---|---|---|---|---|---|---|
| | Treated | Control | Treated | Control | Treated | Control |
| **AFFIRM:** natalizumab vs placebo | 0.23 | 0.73 | 45.6% | 83.1% | 28.2% | 81.2% |
| **ASSESS:** fingolimod vs glatiramer acetate | 0.15 | 0.26 | 44.0% | 79.8% | - | - |
| **CLARITY:** cladribine vs placebo | 0.14 | 0.33 | - | - | 28.9% | 64.1% |
| **CONFIRM:** DMF vs placebo | 0.22 | 0.40 | 81.2% | 82.0% | 67.9% | 87.6% |
| **CONFIRM:** DMF vs glatiramer acetate | 0.22 | 0.29 | 81.2% | 80.4% | 67.9% | 72.1% |
| **DEFINE:** DMF vs placebo | 0.17 | 0.36 | 80.4% | 81.2% | 50.8% | 80.8% |
| **FREEDOMS:** fingolimod vs placebo | 0.18 | 0.4 | 45.1% | 82.2% | - | - |
| **OPERA:** ocrelizumab vs interferon | 0.16 | 0.29 | 25.3% | 74.4% | - | - |
| **TEMSO:** teriflunomide vs placebo | 0.37 | 0.54 | 69.5% | 81.6% | - | - |
| **TENERE:** teriflunomide vs interferon | 0.26 | 0.22 | 71.1% | 74.7% | - | - |
| **TOWER:** teriflunomide vs placebo | 0.32 | 0.5 | 69.7% | 81.8% | - | - |
| **TRANSFORMS:** fingolimod vs interferon | 0.16 | 0.33 | 46.0% | 74.6% | - | - |

*AD: aggregated data; DMF: dimethyl fumarate; EDSS: expanded disability status scale; IPD: individual patient data; RAM: relapse with active MRI; RCT: randomized clinical trial*

**eTable 4. Estimated treatment effect on ACES rate**

| Trial | AD approach | | IPD approach | |
|---|---|---|---|---|
| | **Relative ACES rate (95% CI)** | **P-value** | **Relative ACES rate (95% CI)** | **P-value** |
| **AFFIRM:** natalizumab vs placebo | 1.06 (0.48-2.02) | 0.99 | 1.30 (0.99-1.72) | 0.06 |
| **ASSESS:** fingolimod vs glatiramer acetate | 1.84 (0.56-4.52) | 0.39 | - | |
| **CLARITY:** cladribine vs placebo | - | | 0.76 (0.56-1.04) | 0.09 |
| **CONFIRM:** DMF vs placebo | 0.65 (0.19-1.58) | 0.29 | 1.46 (0.70-3.06) | 0.31 |
| **CONFIRM:** DMF vs glatiramer acetate | 0.85 (0.24-2.19) | 0.58 | 0.72 (0.39-1.35) | 0.31 |
| **DEFINE:** DMF vs placebo | 0.56 (0.16-1.40) | 0.19 | 1.23 (0.71-2.11) | 0.46 |
| **FREEDOMS:** fingolimod vs placebo | 1.48 (0.63-2.96) | 0.42 | - | |
| **OPERA:** ocrelizumab vs interferon | 1.72 (0.71-3.70) | 0.30 | - | |
| **TEMSO:** teriflunomide vs placebo | 1.26 (0.45-2.81) | 0.79 | - | |
| **TENERE:** teriflunomide vs interferon | 1.64 (0.32-6.68) | 0.66 | - | - |
| **TOWER:** teriflunomide vs placebo | 1.17 (0.42-2.56) | 0.92 | - | - |
| **TRANSFORMS:** fingolimod vs interferon | 1.15 (0.43-2.65) | 0.99 | - | - |

*ACES: acute clinical event with stable MRI; AD: aggregated data; DMF: dimethyl fumarate; IPD: individual patient data*

### SUPPLEMENTARY FIGURE

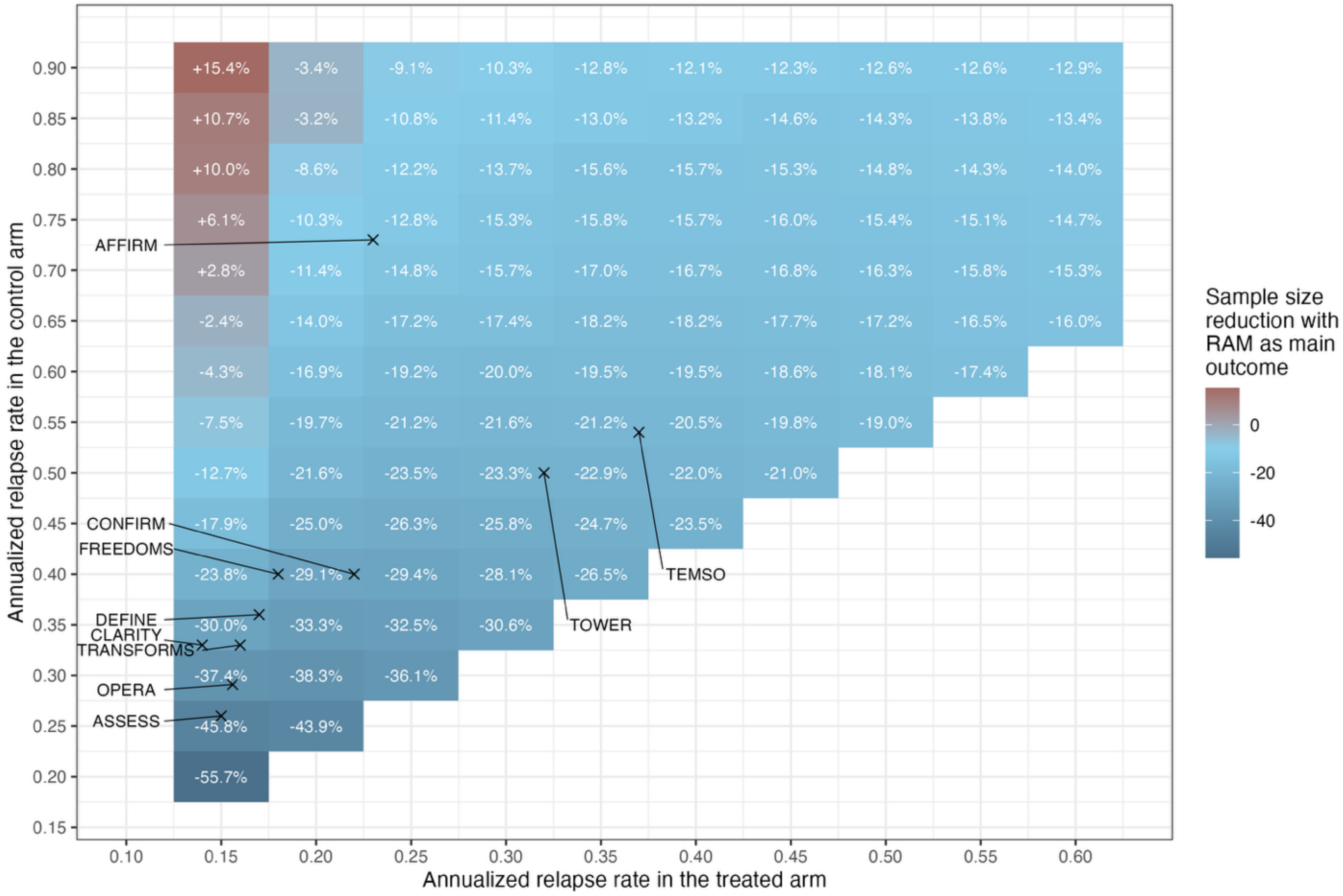


**eFigure 1. Sample size reduction with RAM as the primary outcome compared to clinically defined relapses, with varying ARR in the treated and control arms**

To achieve a statistical power of 90%, with a 5% two-sided alpha risk, with an ACES rate of 0.10/year in each arm.

*ACES: acute clinical event with stable MRI; ARR: annualized relapse rate, RAM: relapse with active MRI*
